# Infectiousness of SARS-CoV-2 breakthrough infections and reinfections during the Omicron wave

**DOI:** 10.1101/2022.08.08.22278547

**Authors:** Sophia T. Tan, Ada T. Kwan, Isabel Rodríguez-Barraquer, Benjamin J. Singer, Hailey J. Park, Joseph A. Lewnard, David Sears, Nathan C. Lo

## Abstract

SARS-CoV-2 breakthrough infections in vaccinated individuals and reinfections among previously infected individuals have become increasingly common. Such infections highlight a broader need to understand the contribution of vaccination, including booster doses, and natural immunity to the infectiousness of persons with SARS-CoV-2 infections, especially in high-risk populations with intense transmission such as prisons. Here, we show that both vaccine-derived and naturally acquired immunity independently reduce the infectiousness of persons with Omicron variant SARS-CoV-2 infections in a prison setting. Analyzing SARS-CoV-2 surveillance data from December 2021 to May 2022 across 35 California state prisons with a predominately male population, we estimate that unvaccinated Omicron cases had a 36% (95% confidence interval (CI): 31-42%) risk of transmitting infection to close contacts, as compared to 28% (25-31%) risk among vaccinated cases. In adjusted analyses, we estimated that any vaccination, prior infection alone, and both vaccination and prior infection reduced an index case’s risk of transmitting infection by 22% (6-36%), 23% (3-39%) and 40% (20-55%), respectively. Receipt of booster doses and more recent vaccination further reduced infectiousness among vaccinated cases. These findings suggest that although vaccinated and/or previously infected individuals remain highly infectious upon SARS-CoV-2 infection in this prison setting, their infectiousness is reduced compared to individuals without any history of vaccination or infection, underscoring some benefit of vaccination to reduce but not eliminate transmission.

## Introduction

Transmission dynamics of SARS-CoV-2 have shifted over the course of the pandemic due to widespread vaccination, natural infection, and emergence of novel variants *(1)*. While the early pandemic was characterized by infections in susceptible individuals, SARS-CoV-2 breakthrough infections among vaccinated individuals and reinfections among previously infected individuals are now increasingly frequent (*2–4*). Following the emergence of the highly infectious Omicron variant in December 2021, the United States observed the largest surge in COVID-19 cases to date (*5*). Determining the impact of vaccination, including booster doses, and prior infection on the infectiousness of persons with Omicron variant infections remains necessary to understand transmission dynamics of these variants.

There is limited data on the infectiousness of breakthrough SARS-CoV-2 infections in vaccinated persons and reinfections with the Omicron variant, especially in high-risk transmission settings such as prisons. Prior to the Omicron variant, available data on the infectiousness of SARS-CoV-2 breakthrough infections in vaccinated individuals was mixed; there was evidence to support reduced infectiousness of breakthrough infections in household studies and through study of viral kinetics (*6-8*), though other studies have found no difference in the infectiousness of primary and breakthrough SARS-CoV-2 infections (*9,10*). This data was predominately among persons immunized only with primary series doses (*6–10*). More recent data from household studies of Omicron variant transmission support that vaccination may reduce infectiousness, although are often limited in capturing detailed aspects of the transmission environment and accounting for interaction with prior infection (*11–13*).

Studying the transmission dynamics of the SARS-CoV-2 Omicron variant and the impact of vaccination and prior infection is especially important in vulnerable, high-risk populations with intense ongoing transmission, such as the incarcerated population. The COVID-19 pandemic has disproportionately affected incarcerated individuals (*14,15*), as transmission of SARS-CoV-2 remains high in prison settings, fueled in part by overcrowding, poor or lack of ventilation, and introduction from community sources despite high vaccination rates among residents (*14,16–21*). In this study, we report on the infectiousness of SARS-CoV-2 infections occurring in vaccinated persons and/or those with prior infection relative to unvaccinated and previously uninfected individuals who were incarcerated in a U.S. state prison system during the first 5 months of the Omicron wave (subvariants BA.1 and BA.2). Our findings have broad implications for public health policy, with particular relevance to incarcerated populations and other high-density congregate living environments.

## Results

### SARS-CoV-2 infections and testing within the study population

We analyzed detailed records of SARS-CoV-2 infection and housing data from all 35 adult institutions in California’s state prison system during periods of high-volume testing, assessing risk of transmission between individuals sharing a cell with solid doors and walls. We aimed to assess the infectiousness of Omicron variant SARS-CoV-2 infections in confirmed index cases, stratified by their vaccine status and prior infection history. We analyzed data during a 5-month interval (December 15, 2021 - May 23, 2022) of widespread circulation of Omicron variants (subvariants BA.1 and BA.2), during periods of both systematic and reactive SARS-CoV-2 testing. In total, there were 22,334 confirmed SARS-CoV-2 infections and 31 hospitalizations due to COVID-19 in the study population (N=111,687) during the study period (Figs. 1-2 and Supplementary Fig. 1). The study population was 97% male based on the population incarcerated in these institutions, and residents were tested on average 8.1 times (interquartile range (IQR): 4-11) for SARS-CoV-2 over the 5-month period. The average time between tests in the study population was 11.7 days (IQR: 4-10) (Supplementary Fig. 2). Most index cases were moved into quarantine or isolation within 3 days of their first positive test. Additional details on testing, quarantine, and isolation in California state prisons are included in Supplementary Notes.

**Figure 1:**
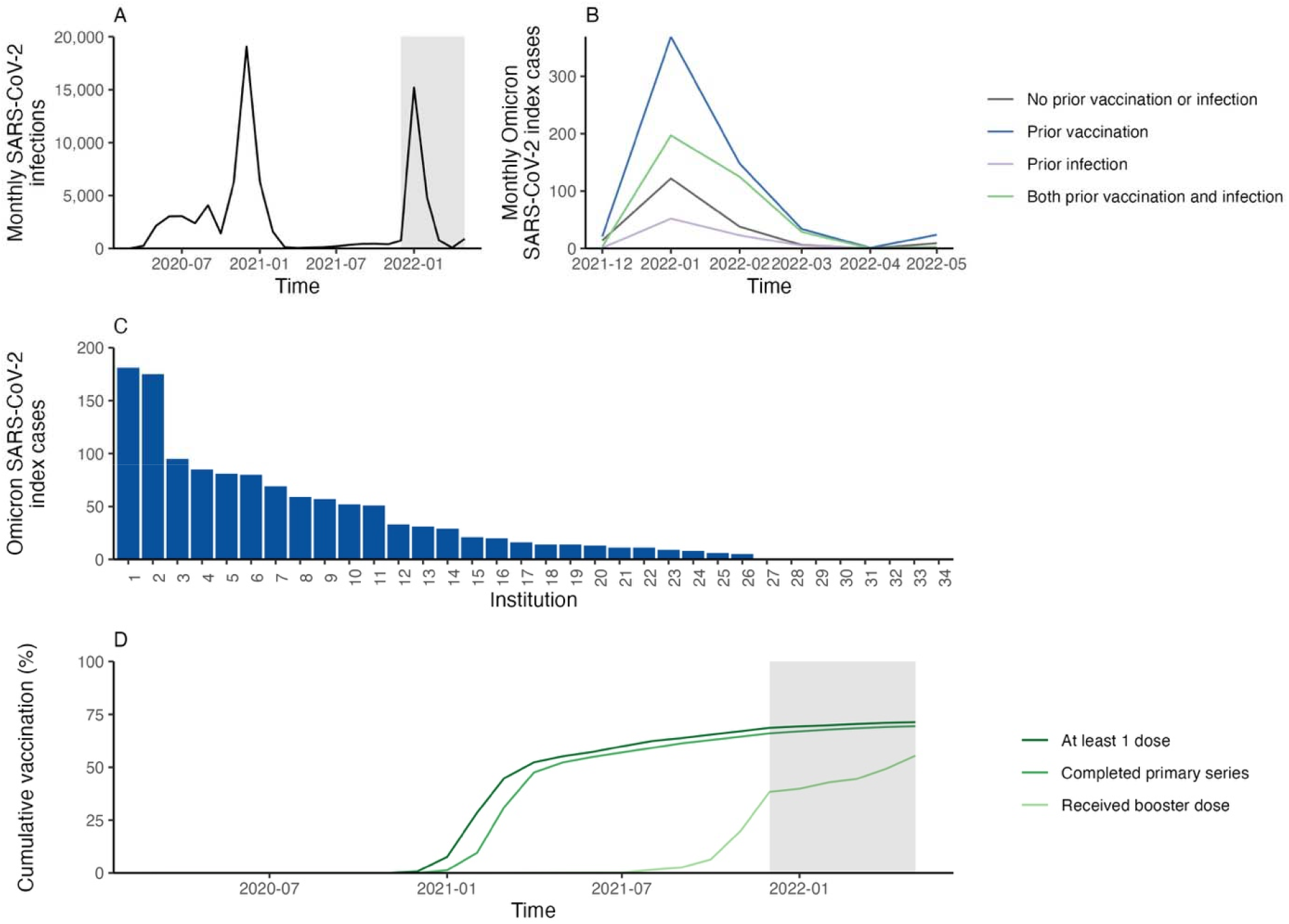
SARS-CoV-2 infections and vaccination over time in the study population in California state prisons. We obtained data on SARS-CoV-2 infections, vaccination, and contact history for residents incarcerated in the California state prison system from March 1, 2020, to May 20, 2022. Panel A shows the number of SARS-CoV-2 infections over time in the study population. Panel B shows the number of SARS-CoV-2 index cases included in the analysis over time, stratified by history of prior natural infection and vaccination. Panel C shows the number of SARS-CoV-2 index cases by institution during the Omicron wave (December 15, 2021, to May 20, 2022) included in the analysis. Panel D shows the COVID-19 vaccine coverage over time for residents in the California state prison system by primary series and booster dose. The shaded region in panels A and D corresponds with the Omicron variant wave.

**Figure 2:**
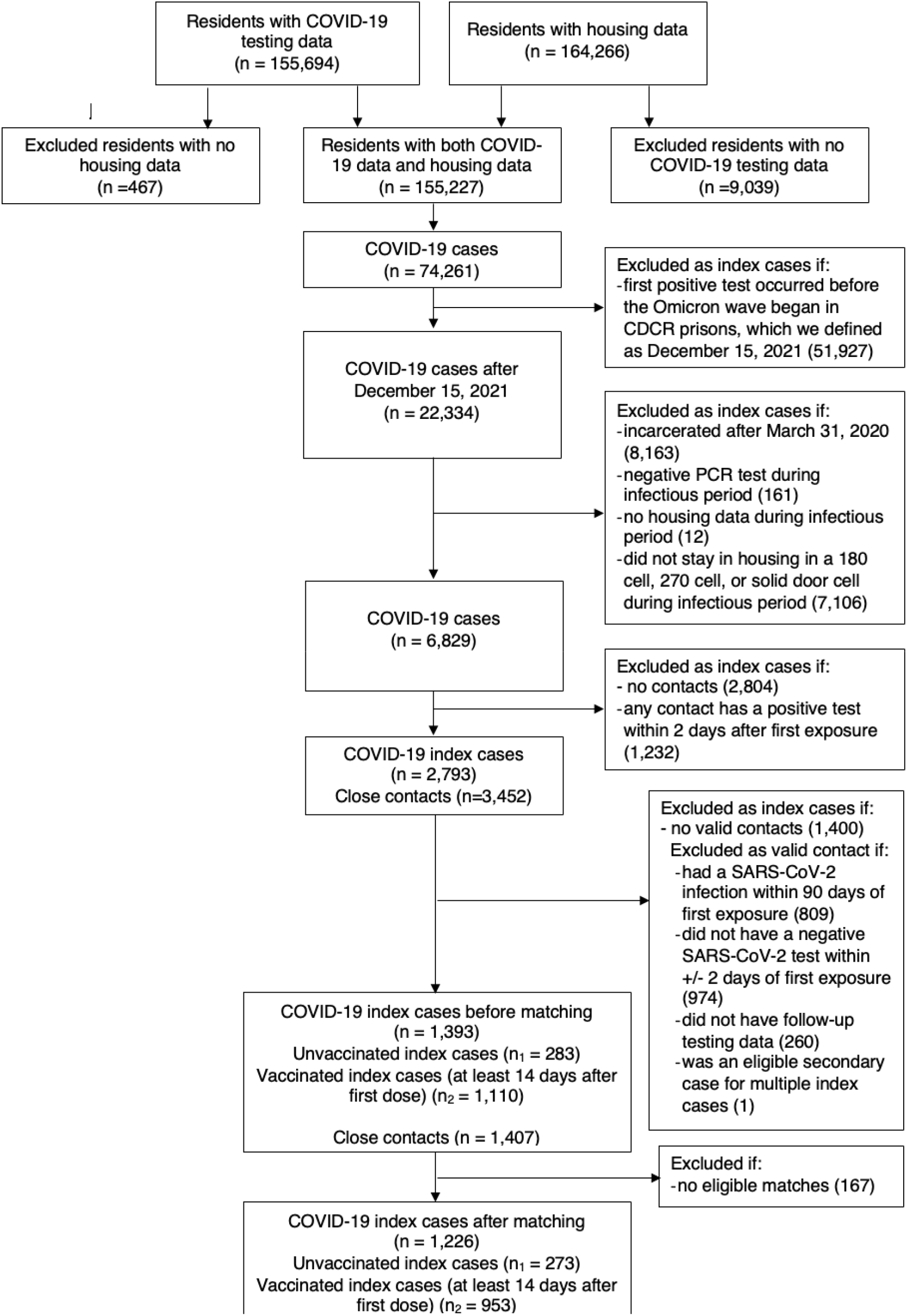
Study population flow chart. We obtained data on residents incarcerated in the California state prison system from March 1, 2020, to May 20, 2022, who were diagnosed with COVID-19 based on a positive molecular test. We applied the inclusion and exclusion criteria to Omicron index cases of COVID-19 and close contacts who shared a cell for at least one night. The sample size at each step is plotted in the figure.

We identified 1,226 index cases over the study period based on the inclusion criteria of having a positive SARS-CoV-2 diagnostic test (without a prior positive test in the preceding 90 days), continuous incarceration beginning prior to April 1, 2020 (to ensure reliable reporting of prior SARS-CoV-2 infection), and a valid close contact in a shared, closed-door cell (Figs. 1-2 and Supplementary Fig. 1). We defined close contacts of the index case as residents who shared a cell with an index case for at least one night while the index case was infectious (assuming a 5-day infectious period following a positive test (*22*)); we required the close contact to have a negative SARS-CoV-2 test within 2 days of first exposure as well as follow up testing data within 14 days after last exposure (64% of close contacts met both criteria). Each index case was assigned a single close contact at random if multiple contacts were identified (<0.1% of cases). Further description of inclusion criteria and exclusion criteria that were needed to address concerns for confounding and misclassification is available in the Methods.

We matched unvaccinated index cases (N=273) and vaccinated index cases (N=953) by institution (exactly) and time (within 30 days) and by a propensity score (for receipt of vaccination), excluding cases without eligible matches (Fig. 2). We matched an average of 3.5 (interquartile range: 2-4) vaccinated index cases to each unvaccinated index case (Supplementary Fig. 3). We observed good balance across matched index cases (Supplementary Figs. 4-5). The mean duration of exposure of close contacts to index cases was 2.4 days for unvaccinated index cases and 2.2 days for vaccinated index cases (Supplementary Fig. 6). The average duration from a close contact’s first exposure to subsequent testing for contacts exposed to a vaccinated and unvaccinated index case was both 6.2 days, and the mean duration of last eligible follow up testing in close contacts occurred at day 10 after first exposure for unvaccinated index cases and 10.6 days for vaccinated index cases (Supplementary Fig. 7). The distribution of secondary cases from time since exposure was similar between vaccinated and unvaccinated index cases (6.7 versus 5.7 days, Supplementary Fig. 8). Descriptive data on the study population’s demographics, vaccine uptake, and prior infections are shown in Table 1 and Supplementary Table 1.

**Table 1:** Characteristics of study population of COVID-19 index cases and close contacts in California prisons.

|  | Index cases (N=1226)<br>(N (%) or mean (SD)) |  | Close contacts * (N=1226)<br>(N (%) or mean (SD)) |  |
| --- | --- | --- | --- | --- |
|  | No COVID-19<br>vaccination<br>(N=273) | Any COVID-19<br>vaccination (N=953) | No COVID-19<br>vaccination<br>(N=173) | Any COVID-19<br>vaccination<br>(N=1053) |
| **Sex** |  |  |  |  |
| Female | 8 (3%) | 30 (3%) | 7 (4%) | 31 (3%) |
| Male | 265 (97%) | 923 (97%) | 166 (96%) | 1022 (97%) |
| **Age (years)** | 36.3 (10) | 39 (10.7) | 35.9 (10.1) | 39.6 (11.1) |
| **Race/ethnicity** |  |  |  |  |
| American Indian/Alaskan Native | 0 (0%) | 11 (1%) | 1 (1%) | 10 (1%) |
| Asian or Pacific Islander | 4 (2%) | 12 (1%) | 0 (0%) | 10 (1%) |
| Black | 89 (33%) | 221 (23%) | 56 (32%) | 244 (23%) |
| Hispanic | 145 (53%) | 548 (58%) | 88 (51%) | 599 (57%) |
| White | 28 (10%) | 136 (14%) | 23 (13%) | 155 (15%) |
| Other ** | 7 (3%) | 25 (3%) | 5 (3%) | 35 (3%) |
| **COVID-19 risk score (range 0-12) \*\*\*** | 0.7 (1.3) | 1 (1.4) | 0.7 (1) | 1.1 (1.5) |
| **Number of days of exposure between index case and close contact** | 2.4 (1.2) | 2.2 (1.1) | 2.4 (1.2) | 2.3 (1.1) |
| **Prior infection** | 84 (31%) | 356 (37%) | 59 (34%) | 448 (43%) |
| **Vaccination status** |  |  |  |  |
| *Unvaccinated* | 273 (100%) | - | 173 (100%) | - |
| *Ad26.COV2* | - | 113 (12%) | - | 152 (15%) |
| Completed only primary series | - | 58 (51%) | - | 70 (46%) |
| Received booster or additional doses | - | 55 (49%) | - | 82 (54%) |
| *BNT162b2* | - | 188 (20%) | - | 193 (18%) |
| Received 1 dose of primary series | - | 5 (3%) | - | 3 (2%) |
| Completed only primary series | - | 55 (29%) | - | 43 (22%) |
| Received booster or additional doses | - | 128 (68%) | - | 147 (76%) |
| *mRNA-1273* | - | 652 (68%) | - | 708 (67%) |
| Received 1 dose of primary series | - | 13 (2%) | - | 25 (4%) |
| Completed only primary series | - | 229 (35%) | - | 223 (31%) |
| Received booster or additional doses | - | 412 (63%) | - | 462 (65%) |

History of prior natural infection and vaccination status reflect the index case and close contact's vaccination and natural infection status on the day of first positive test (for index cases) or first exposure to an infectious index case (for close contacts)
\*Residents were considered close contacts of two index cases in rare cases (<0.01% of cases)
\*\*Other race/ethnicity based on self-reported data and those with mixed race/ethnicity
\*\*\*COVID-19 risk score was estimated by California Correctional Health Care Services as weighted sum of different comorbidities most associated with severe COVID-19 complications (<sup>37</sup>)

### Infectiousness of SARS-CoV-2 breakthrough- and re-infections

Over an average 2.3 days of exposure to the index case, the unadjusted risk of transmission to all close contacts of index cases was 30% (95% CI: 27-32%). Unvaccinated index cases had a 36% (31-42%) risk of transmitting to close contacts, while vaccinated index cases had a 28% (25-31%) risk of transmitting to close contacts (Fig. 3). Index cases with a history of prior SARS-CoV-2 infection (i.e., reinfection) had a lower risk of transmitting to close contacts [23% (19-27%)] than index cases with no history of prior infection [33% (30-37%)]; reduced risk of transmission from index cases who were previously infected was apparent in strata of index cases who had or had not been vaccinated, and who did or did not receive a booster dose (Fig. 3 and Supplementary Table 2).

**Figure 3:**
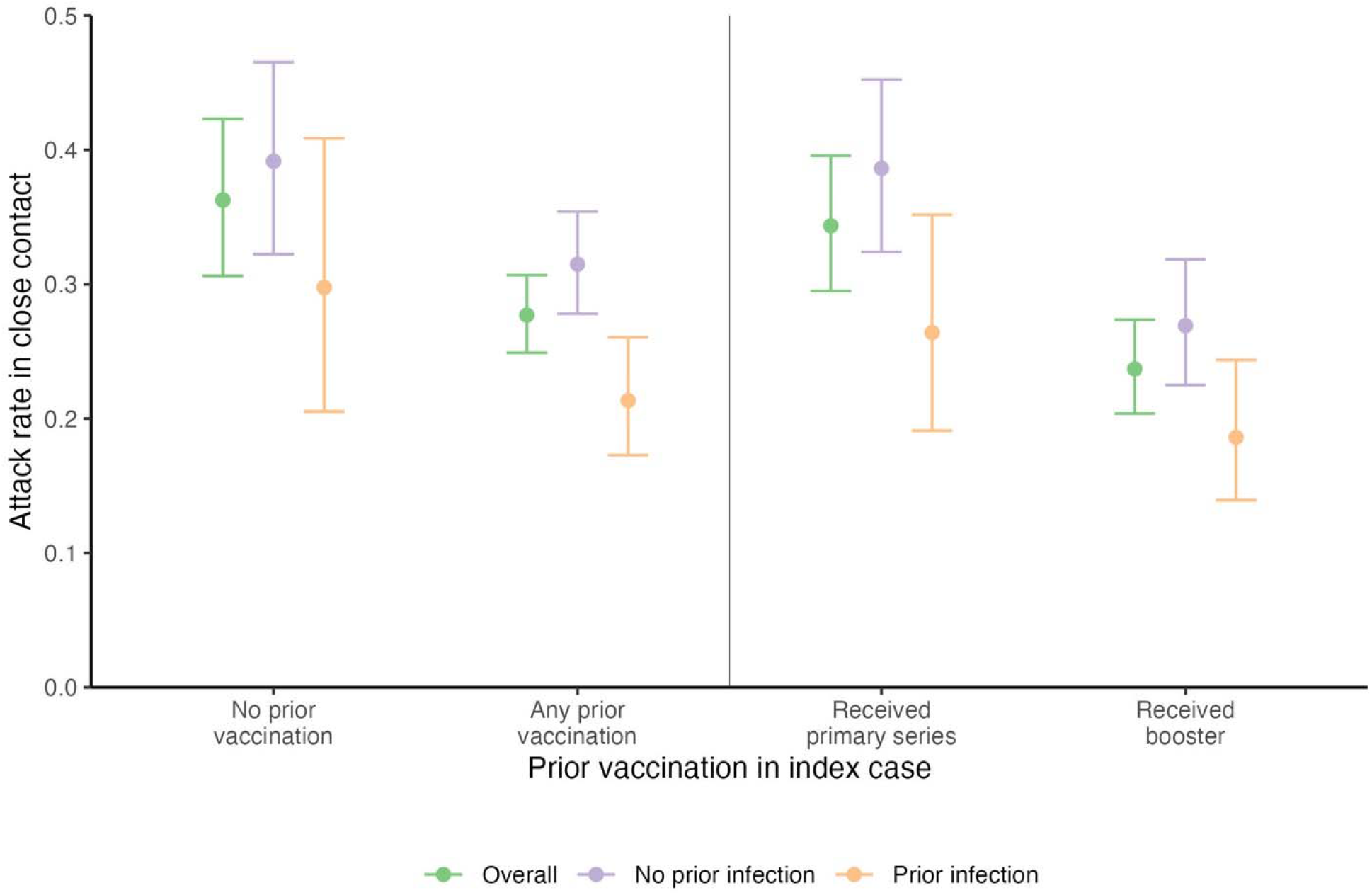
Unadjusted Omicron SARS-CoV-2 attack rate in close contact based on index cases’ vaccine and prior natural infection status. We identified index cases of SARS-CoV-2 infections in residents of the California state prison system who were in close contact with another resident who was confirmed negative for SARS-CoV-2 at the time of contact. We estimated the outcome of subsequent SARS-CoV-2 infection in the close contact under different immune conditions of the index case, with a composite study outcome of attack rate. The attack rate is the probability of infection in the close contact given exposure to an index case. We plot the unadjusted attack rate (represented by points) and 95% confidence intervals (represented by error bars) of SARS-CoV-2 in the close contact stratified by the index cases’ overall vaccine status, the number of vaccine doses in the index case, and index cases’ history of natural infection.

Adjusting for duration of exposure between index cases and close contacts, close contacts’ history of vaccination and prior infection, and facility effects and background SARS-CoV-2 incidence via a robust Poisson regression model, we estimated that index cases who had received ≥1 COVID-19 vaccine doses had 22% (6-36%) lower risk of transmitting infection than unvaccinated index cases. In analyses that further accounted for the number of vaccine doses received by an index case, each additional dose was associated with an average 11% reduction (5-17%) in risk of transmission to the close contact (Fig. 4 and Supplementary Tables 3-5). Prior SARS-CoV-2 infection was similarly associated with a 23% reduction (3-39%) in risk of transmission from the index case. Having both prior vaccination and SARS-CoV-2 infection was associated with a 40% (20-55%) reduction risk of transmission by the index case, based on a linear combination of regression coefficients (Fig. 4); we did not identify evidence of interaction between prior vaccination and prior natural infection associated with transmission risk (Supplementary Table 6).

**Figure 4:**
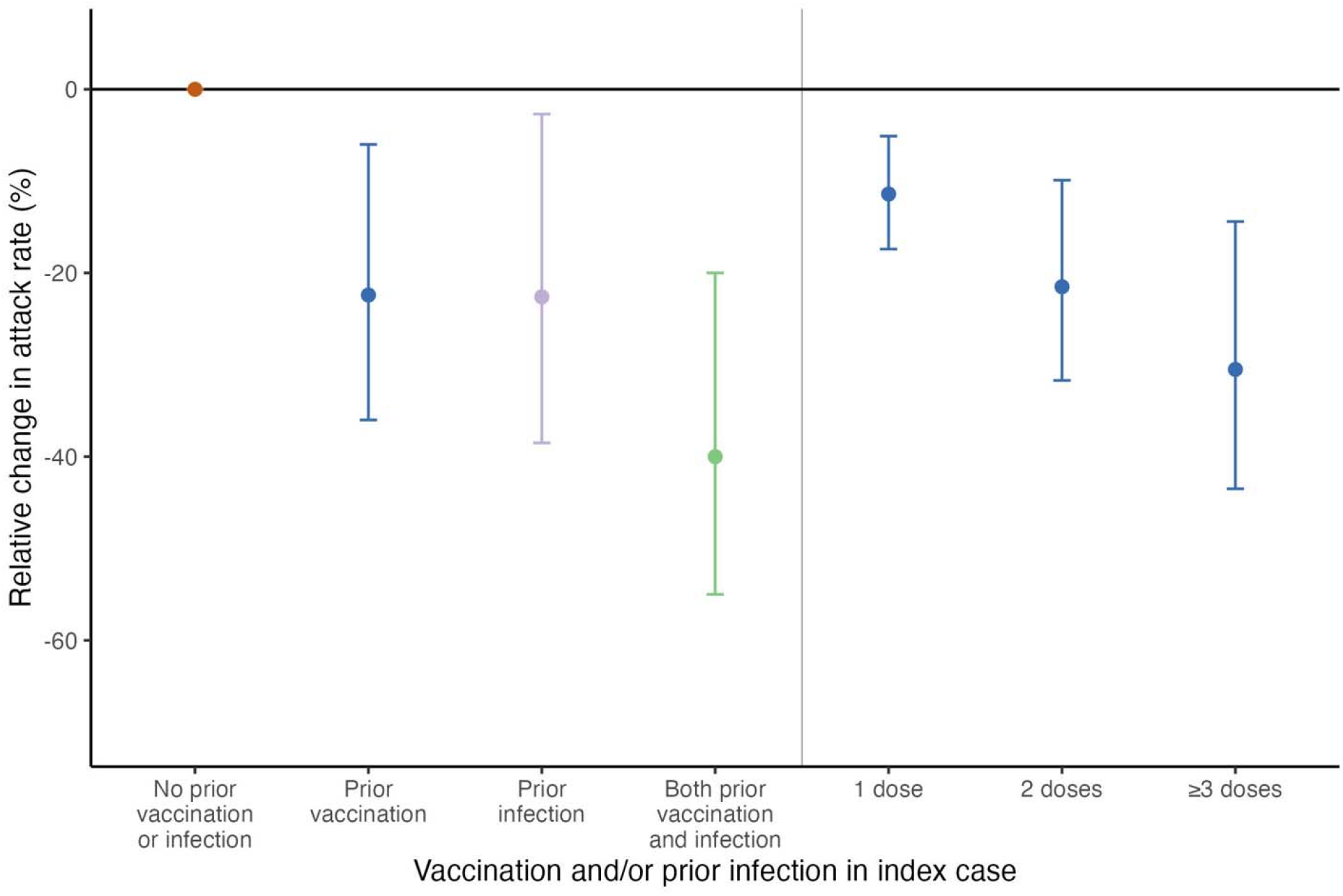
Relative change in Omicron SARS-CoV-2 attack rate in close contacts based on index cases’ vaccine and prior natural infection status in an adjusted model. We applied a robust Poisson regression model to estimate the relationship between vaccination and natural immunity in index cases on their risk of SARS-CoV-2 transmission in close contacts. We plotted the adjusted relative reduction in infectiousness of index cases (represented as points), as measured via attack rate in close contacts, conferred by vaccination alone, prior infection alone, and both prior vaccination and infection. The estimate for both prior vaccination and infection is based on a linear combination of regression coefficients, given lack of formal statistical interaction between vaccination and prior infection. We conducted a separate regression analysis (right side of graph) that was stratified based on the number of vaccine doses received by the index case. We plotted cluster-robust 95% confidence intervals (represented by error bars).

We assessed the association between time since last vaccine dose and/or natural infection on infectiousness of a SARS-CoV-2 infection and found that time since last dose of a COVID-19 vaccine (as a continuous variable) was associated with increased infectiousness of SARS-CoV-2 infections; for every 5 additional weeks since last vaccine dose, SARS-CoV-2 breakthrough infections were 6% (2-11%) more likely to transmit infection to close contacts. We did not observe a statistically significant relationship between time since last SARS-CoV-2 infection and risk of transmission (Supplementary Table 7 and Supplementary Fig. 9).

We conducted a number of sensitivity and additional model analyses to evaluate the robustness of the study findings. We evaluated primary study outcomes when relaxing exclusion criteria for close contacts; any prior COVID-19 vaccination was associated with 23% (8-35%) reduction in attack rate when we included close contacts that tested positive within two days of exposure to the first index case and 19% (3-33%) reduction when we removed the requirement of a negative test in close contacts within two days of first exposure to an index case (Supplementary Table 8). Study findings were also similar across changes in the matching process (Supplementary Table 9). Varying definitions of the start and duration of the infectious period attenuated some of the findings (Supplementary Table 10). We found excluding index cases that received the *Ad26*.*COV2* vaccine due to its reduced effectiveness compared to mRNA vaccines led to similar results (Supplementary Table 11). We repeated the primary adjusted analysis using a logistic regression model and found that both prior vaccination [odds ratio (OR) 0.66 (0.48-0.91)] and prior infection [OR 0.68 (0.49-0.95)] were associated with reduced odds of infection in close contacts (Supplementary Table 12). Additional details on sensitivity analyses are available in the Methods.

### Transmission from primary-, breakthrough-, and re-infections

We estimated that primary infections (15% of index cases) contributed to 20% (16-25%) of transmission to secondary cases, breakthrough infections (49% of index cases) contributed to 52% (47-57%) of transmission to secondary cases, reinfections (7% of index cases) contributed to 7% (5-10%) of transmission to secondary cases, and breakthrough infections in previously infected residents (29% of index cases) contributed to 21% (17-26%) of transmission to secondary cases in the study population. We observed similar results over the entire study period (Supplementary Table 13).

## Discussion

Using detailed epidemiologic data from SARS-CoV-2 surveillance within the California state prison system, we found that vaccination and prior infection reduced the infectiousness of SARS-CoV-2 infections during an Omicron-predominant (subvariants BA.1 and BA.2) period. Vaccination and prior infection were each associated with comparable reductions in infectiousness during SARS-CoV-2 infection, and notably, additional doses of vaccination (e.g., booster doses) against SARS-CoV-2 and more recent vaccination led to greater reductions in infectiousness. Of note, reductions in transmission risk associated with prior vaccination and infection were found to be additive, indicating an increased benefit conferred by vaccination for reducing cases’ infectiousness even after prior infection. Irrespective of vaccination and/or prior natural infection, SARS-CoV-2 breakthrough infections and reinfections remained highly infectious and were responsible for 80% of transmission observed in the study population, which has high levels of both prior infection and vaccination. This observation underscores that vaccination and prevalent naturally acquired immunity alone will not eliminate risk of SARS-CoV-2 infection, especially in higher risk settings such as prisons.

Prior studies during the Delta variant wave and prior to widespread booster vaccination are mixed on whether SARS-CoV-2 breakthrough infections in vaccinated persons are potentially less infectious (*6–8*) or equally infectious (*9,10*) to primary infections. In more recent household contact studies during the Omicron variant era (*11–13*), vaccination often led to reduced SARS-CoV-2 infectiousness. Several factors may have enhanced our ability to observe statistically meaningful findings in the present study. The risk of transmission among close contacts in the prison setting and consistency in contact structure, especially in light of increased transmissibility of the Omicron SARS-CoV-2 variant, may have enhanced statistical power in our sample. Relatedly, a higher proportion of index cases in our sample were previously vaccinated or infected, further enhancing the opportunity to compare transmission risk from vaccinated or unvaccinated index cases, and from those who were previously infected or previously uninfected.

A key result is that the vaccine-mediated reduction in infectiousness of SARS-CoV-2 breakthrough infections appears to be dose dependent. Each dose of the vaccine provided an additional average 11% relative reduction in infectiousness, which was mostly driven by residents with a booster dose. The findings of this study support the indirect effects of COVID-19 vaccination (especially booster doses) to slow transmission of SARS-CoV-2 and build on evidence of the direct effects of COVID-19 vaccination (*23*) to emphasize the overall importance of COVID-19 vaccination. The public health implication of these findings is further support for existing policy using booster doses of vaccination (*24*) to achieve the goal of lowering population level transmission. The impact of additional bivalent vaccine doses, which are now authorized for persons over 5-6 years old (*25*), on transmission should be a priority for further study. Additional considerations about the timeliness of vaccine doses are also necessary as we found that index cases with more distant history of COVID-19 vaccination had higher risk of transmission of infection to close contacts. Given this finding, this study raises the possibility of timed mass vaccination in incarcerated settings during surges to slow transmission.

The findings from this study have direct implications in addressing COVID-19 inequities in the incarcerated population through additional vaccination. In California state prisons at time of the study, although 81% of residents and 73% of staff have completed a primary vaccination series, only 59% of residents and 41% of staff have received the number of vaccination doses recommended by the Centers for Disease Control and Prevention based on their age and comorbid medical conditions (*26*). Our findings also provide a basis for additional considerations for housing situations of cases based on prior vaccination and infection history in future surges and can be used alongside other measures, such as depopulation and ventilation interventions, to protect incarcerated populations.

However, this study also underscores the persisting vulnerability to COVID-19 among residents and staff in correctional settings despite widespread vaccination, natural immunity, and use of non-pharmaceutical interventions. The overall attack rate of SARS-CoV-2 among cellmates in the study population (who were generally moved into isolation following symptoms or a positive test) was 30%, and index cases with breakthrough infections or reinfections remained highly infectious, which call into question the ability of high vaccination rates alone to prevent all SARS-CoV-2 transmission in correctional settings. In the United States, which incarcerates more residents per capita than any other country in the world (*26*) and has a quarter of the world’s incarcerated population, correctional settings are characterized by poorly ventilated facilities, populations with increased rates of comorbid health conditions, high-risk dormitory housing, and overcrowding (*18,27–29*). Given the inability of current efforts to reduce transmission of SARS-CoV-2, decarceration efforts may be the most likely to have substantial effects on reducing cases.

The secondary attack rate in this study was on the lower end of published estimates when comparing to household studies. Of note, the secondary attack rate of the SARS-CoV-2 Omicron variant in recent household studies ranges from 29-53% (*11–13*), in contrast to a 30% attack rate in this study. The prison environment has distinct epidemiologic differences to households. The dense living environment increases the likelihood of transmission in the prison environment compared to a household, while the frequent asymptomatic testing (with isolation of positive cases) in the prisons likely reduced the exposure time and subsequent transmission risk compared to households. The transmission of the prison cell is also likely more uniform than a household.

Strengths of this study include access to detailed records of all residents in the California state prison system, encompassing individuals’ prior COVID-19 vaccine receipt and prior natural infection history (based on frequent testing throughout the pandemic), as well as social network given record of where residents slept each night over the study period. We use a consistent definition of social contact between the index case of COVID-19 and close contact based on the uniformity of cell type. The frequent testing ensures early identification of infections and systematic capture of asymptomatic and symptomatic infections to avoid bias by participants’ immune status (which could affect temporal onset of symptoms). The risk of misclassification of close contacts is low given most follow up testing in close contacts occurred well after first exposure to an index case (Supplementary Notes). The large sample size facilitates analyses of the contribution of combinations of prior vaccination statuses and natural infection on risk of transmission, including analyses examining the impact of booster doses.

Limitations should also be considered. We cannot exclude the possibility of some residual confounding (e.g., behavioral differences that affect risk of transmission) between persons who were vaccinated against SARS-CoV-2 and those who were unvaccinated. There is a possibility that close contacts who test positive for SARS-CoV-2 were not infected by their assigned index case but instead by interaction with infectious individuals outside of their cell. However, this misattribution would be expected to dampen apparent associations of transmission risk with index cases’ vaccination status and infection history, but not bias the relative estimates. To further address the risk of misattribution, we adjust for background SARS-CoV-2 incidence and match contact pairs by facility and time. Our study population is a subset of the entire incarcerated population in California and may not represent all incarcerated settings. Studies of SARS-CoV-2 infectiousness may be subject to biases (*30–32*). The strict inclusion and exclusion criteria in this study may introduce bias into the analysis, although we perform sensitivity analyses on these criteria with overall consistent findings. We also adjust for prior infection in analyses to account for potential concerns about differential susceptibility related to prior infection in vaccinated versus unvaccinated persons. Given limited SARS-CoV-2 testing capacity early in the pandemic and some residents’ decision to decline testing, it is possible infections among some residents may not have been captured, although such misclassification would be expected to bias our findings to the null. SARS-CoV-2 testing was variable over time in the prison system, with periods of routine weekly testing and other periods of reactive testing; however, periods without reactive testing align with times during which SARS-CoV-2 was unlikely to be circulating at high levels within the facilities, suggesting this is unlikely to bias results substantially. The study findings on boosters may also be related to recent vaccination effects. This study design did not provide a basis for identifying effects of vaccination and prior infection on risk of acquiring SARS-CoV-2 among close contacts, although we did adjust for prior infection and vaccination in close contacts in the primary analysis. Of note, vaccine effectiveness against infection among incarcerated persons has been reported within this population during earlier periods (*33,34*). We do not have a detailed record of person-level masking, symptoms, cycle thresholds for polymerase chain reaction testing, or serologic testing. During the study, the predominant Omicron subvariants in California and California prisons were BA.1 and BA.2 based on genomic surveillance, although we did not genotype every SARS-CoV-2 isolate in this study.

This study demonstrates that breakthrough COVID-19 infections with the Omicron variant remain highly infectious, but that both vaccination and natural infection confer reductions in transmission, with benefit of additional vaccine doses. As SARS-CoV-2 breakthrough infections and reinfections become the predominant COVID-19 case, this study supports the importance of booster doses in reducing population level transmission with consideration of mass timed vaccination during surges, with particular relevance in vulnerable, high-density congregate settings.

## Methods

### Data

We used data from the California Correctional Health Care Services (CCHCS), which included anonymized person-level data on SARS-CoV-2 testing, COVID-19 vaccination, and nightly resident housing for incarcerated persons in the California state prison system from March 1, 2020, to May 20, 2022. The objective of the study was to study the relative infectiousness of Omicron SARS-CoV-2 breakthrough infections and reinfections. We defined the period of the Omicron variant wave as between December 15, 2021, and May 20, 2022, based on genomic surveillance data from the California prison system. This project was approved by the institutional review board (IRB) at the University of California, San Francisco (see Ethics and IRB approval).

### COVID-19 index case definition and infectious period

The inclusion and exclusion criteria for an index case of COVID-19 for the study are shown in Figure 2. We defined an index case as a resident with any conclusive positive SARS-CoV-2 diagnostic test. The majority of tests (83%) were polymerase chain reaction. We excluded index cases with a prior positive test within the preceding 90 days (unless they had a negative PCR test in the interim), as well as those with a false positive or inconclusive result. We included only infections that occurred in residents who were incarcerated continuously beginning prior to April 1, 2020, to ensure consistent reporting of prior SARS-CoV-2 infection given that these data are not available from recently incarcerated residents. We classified index cases based on their COVID-19 vaccination status and prior natural infection history. We defined SARS-CoV-2 breakthrough infections as a positive SARS-CoV-2 diagnostic test occurring in persons at least 14 days after their first dose of vaccine, as long as that person did not have a prior positive diagnostic test in the preceding 90 days. We defined reinfection as a positive SARS-CoV-2 diagnostic test occurring in persons with a prior laboratory-confirmed natural infection provided that at least 90 days had elapsed since the first infection unless they had received a negative SARS-CoV-2 PCR test in the interim.

For a conservative measure of the time each index case was infectious, we counted from the date of an index case’s first positive SARS-CoV-2 test through 5 days thereafter (*22,35,36*). We varied the start and the duration of the infectious period in sensitivity analyses. We shortened the assumed 5-day infectious period for a COVID-19 index case if the resident had a negative rapid antigen test during the infectious period. We excluded index cases if the resident had a negative PCR test during the infectious period to mitigate potential bias due to delayed detection of cases. Isolation and quarantine protocols in the prison system are described in the Supplementary Notes.

### Close contacts of COVID-19 index case

We defined a close contact of a COVID-19 index case as any resident who shared a cell with an index case while the index case was considered infectious, per the above definition. We further required a close contact to test negative for SARS-CoV-2 within 2 days before or after first exposure to an index case (to reduce the chance they were already infected by another resident) and to have follow up testing within 3 to 14 days after last exposure. Close contacts were excluded if they had a prior SARS-CoV-2 infection in the preceding 90 days (unless they had a negative PCR test in the interim). Characteristics of close contacts included or excluded due to missing testing data are shown in Supplementary Table 14. We defined first exposure as the first day that the index case and close contact shared a room during the index case’s infectious period (based on a positive test in the index case). To limit misattribution of secondary cases and close contacts, we only included contacts that shared a solid-door cell with fewer than 10 total residents during the index case’s infectious period (>95% of index cases had 3 or fewer persons per cell). The solid-door cell type was chosen to provide a more consistent transmission environment for comparison between facilities and to improve attribution of infection to the index case (versus residents in nearby cells with cell types with open air exchange). We defined secondary SARS-CoV-2 infection as close contacts who tested positive for SARS-CoV-2 between 3 days after first exposure and 14 days after last exposure to the index case. We excluded close contacts that were secondary cases for multiple index cases (only 1 close contact). After other inclusion and exclusion criteria were applied to close contacts, if index cases had more than one valid close contact (<0.1% of index cases and index cases had no more than 3 valid close contacts), we randomly selected a single contact to include in the analysis.

### Statistical analysis

We performed matching of unvaccinated index cases and vaccinated index cases to limit confounding and to account for heterogeneity of SARS-CoV-2 epidemiology across institutions and over time. We first estimated the propensity for index cases to receive vaccination using logistic regression based on their age, prior history of SARS-CoV-2 infection, and COVID-19 risk score based on co-morbid conditions related to risk of severe disease (*37*). We then applied 1:10 nearest matching on institution (exact), time (< 30 days) and propensity score (caliper choice of 0.1) scaled to be weighted equally and matched without replacement (*38*). We excluded any index cases without matches.

We estimated unadjusted attack rates, defined as the proportion of close contacts who tested positive between 3 days after initial exposure and 14 days after last exposure with an index case, and computed associated 95% binomial confidence intervals (95% CI). We estimated attack rates by number of vaccine doses and prior natural infection.

To estimate the relative infectiousness of SARS-CoV-2 breakthrough infections and/or SARS-CoV-2 reinfections, we fit a Poisson regression model with robust errors to account for key variables in an adjusted analysis. We used Poisson regression in the main analysis since coefficients are easier to interpret than those in logistic regression and may be more robust with model misspecification (*39*). Since binomial data violates distributional assumptions, robust errors were computed. The primary study outcome was binary, the SARS-CoV-2 infection outcome in the close contact. The exposure of interest was the vaccine status (primary analysis with binary vaccine status, alternative analysis with number of vaccine doses) of the index case, which can be interpreted as the relative change in attack rate in the close contact based on the index cases’ vaccine status. We also adjusted for index case’s prior SARS-CoV-2 infection history, duration of exposure between index case and close contact, close contact’s vaccine status (number of doses) and prior natural infection, institution, and institution-specific SARS-CoV-2 incidence in the 7 days leading up to infection in the index case. The regression model accounted for matching weights and cluster-robust standard errors based on matching group membership. We did not use repeated measured data. We did not perform a formal sample size calculation, although the final sample size would be expected to detect a minimum 10% difference between groups. The pre-analysis plan is publicly available (*40*).

We classified secondary infections (N=363) among close contacts by index cases’ prior vaccination and/or infection history and estimated the crude fraction of secondary infections that were attributable to different index cases as well as their respective 95% binomial confidence intervals. We additionally estimated the attributable fraction of transmission among all SARS-CoV-2 infections in the study period. We first estimated the adjusted attack rate of SARS-CoV-2 infection by a case’s prior vaccination and/or infection history using estimates of the relative reduction in infectiousness. We then applied the attack rates to the observed number of infections to estimate the attributable fraction of transmission by prior vaccination and prior infection status. Analysis was conducted in R (version 4.2.1).

### Sensitivity analysis

We conducted an alternative analysis that defined the index case vaccine status by number of doses (rather than binary) and assessed the relationship between number of vaccine doses and risk of secondary infection in close contacts. Matches were reweighted within vaccine groups with confirmation of covariate balance across groups (Supplementary Table 1) (*38*); we also repeated matching to a single reference group to maximize balance. We further examined the relationships between prior vaccination, prior infection, and infectiousness of an index case by testing a formal interaction between prior vaccination and prior natural infection and evaluated the relationship between the time since most recent exposure (as continuous variable) to either COVID-19 vaccination or SARS-CoV-2 infection. We varied definitions of COVID-19 vaccine status in close contacts in sensitivity analyses. We assessed impact of relaxing different exclusion criteria for index cases and close contacts on study results. We varied the start and duration of the infectious period in sensitivity analyses. To assess model robustness, we evaluated study outcomes under different matching specifications and when using a logistic regression model. Given the lower vaccine effectiveness of the *Ad26*.*COV2* vaccine, we conducted a sensitivity analysis removing index cases that received the *Ad26*.*COV2* vaccine.

### Ethics and IRB approval

This project was approved by the IRB at the University of California, San Francisco (IRB #: 21-34030). The IRB waived the need for informed consent given use of secondary datasets deemed no more than minimal risk based on the following criteria: deemed no more than minimal risk to subjects; could not practicably be done without the waiver; could not practicably be done without identifiable information; will not adversely affect rights and welfare of subjects; and will provide subjects with additional pertinent information after participation.

### Data availability

Data requests may be made to the California Correctional Health Care Services and are subject to controlled access due to requirements to enhance protection of this vulnerable incarcerated population. Therefore, de-identified person-level data from this dataset is not publicly available. Requests for data access for study replication or new analyses can be made here: http://cdcrdata.miraheze.org/wiki/Request_data.

### Code availability

All analytic code is publicly available here: https://github.com/sophttan/CDCR-CalProtect.

## Data Availability

Data requests may be made to the California Correctional Health Care Services and are subject to controlled access due to requirements to enhance protection of this vulnerable incarcerated population. Therefore, de-identified person-level data from this dataset is not publicly available. Requests for data access for study replication or new analyses can be made here: http://cdcrdata.miraheze.org/wiki/Request_data. All analytic code is publicly available.

https://github.com/sophttan/CDCR-CalProtect

## Acknowledgments

We thank the individuals at the Office of the California Prison Health Care Receivership and California Correctional Health Care Services, as well as all those involved in the ongoing response to the COVID-19 pandemic in California. We acknowledge the individuals who provided the data underlying these analyses.

## Disclaimer

The study content is solely the responsibility of the authors and does not necessarily represent the official views of the National Institutes of Health.

## Author contributions

Ms. Sophia Tan and Dr. Nathan Lo had full access to all the data in the study and take responsibility for the integrity of the data and the accuracy of the data analysis.

Study concept and design: JAL, IRB, DS, NCL

Statistical analysis: STT, IRB, NCL

Acquisition, analysis, or interpretation of data: All authors

First draft of the manuscript: STT, NCL

Critical revision of the manuscript: All authors

Contributed intellectual material and approved final draft: All authors

