## Supplementary Material for "Infectiousness of SARS-CoV-2 breakthrough infections and reinfections during the Omicron wave"

**Contents**

Supplementary Notes	2

Supplementary Tables	3

Supplementary Figures	10

**Supplementary Notes**

*SARS-CoV-2 testing, quarantine, and isolation practices within California state prisons*

Testing practices for SARS-CoV-2 varied over time within the California state prison system. The majority of tests were polymerase chain reaction (PCR). Residents were tested both reactively, if residents were symptomatic or were considered close contacts of a confirmed COVID-19 case, and systematically (weekly) during periods of high transmission. Following a positive test, most residents were not tested for the following 90 days. The distributions for the number of tests received by residents in the study population and the time between tests over the study period are shown in Supplementary Figure 3.

When residents received a positive SARS-CoV-2 test result, they were moved into isolation for 10-14 days. Isolation included single cell occupancy (provided there was sufficient housing within the prison) or group isolation with other residents with active SARS-CoV-2 infection. Residents generally remained in their assigned housing while their SARS-CoV-2 test results were pending unless they were symptomatic.

*Concern about misclassification of secondary SARS-CoV-2 infections*

To address concerns about misclassification of secondary SARS-CoV-2 infection among close contacts of index cases (specifically testing too early to capture infection), we further examined the frequency and timing of follow up testing among close contacts. Close contacts included in the study population received on average 2 SARS-CoV-2 tests (IQR: 1-2) during the follow up period. Close contacts received their first follow up test 6.2 days (IQR: 4-7) after first exposure to an index case. The last follow up SARS-CoV-2 test among close contacts that received >1 follow up test (68% of close contacts) was on average 12.2 days (IQR: 11-14) after first exposure. The distributions for the number of tests received during follow up and timing of follow up are similar between close contacts of unvaccinated and vaccinated index cases and are shown in Supplementary Figure. 9. Among all close contacts, we found that the timing of the last eligible test in the close contact is 10.6 days after first exposure (IQR: 7-14) for those with a vaccinated index case and is 10 days after first exposure (IQR: 6-14) for those with an unvaccinated index case. Given most testing in close contacts was sufficiently after the exposure period, the frequency of follow up testing, and the balanced timing in follow up testing between vaccinated and unvaccinated index cases, the risk of misclassification is overall low.

*Pre-analysis plan*

The pre-analysis plan is publicly available here: <https://github.com/sophttan/CDCR-CalProtect>*.* The final analysis had minor deviations from the initial plan. We removed the random effects for each index case since the final dataset did not include repeated measures data (<0.1% of dataset, which was removed). We removed the inclusion criteria that index cases must have a negative PCR SARS-CoV-2 test in the 8 days prior to first positive test in order to achieve an adequate sample size. This change would not be expected to have differential implications for eligibility of index cases who were vaccinated and unvaccinated. We increased the number of matches to 1:10 to maximize sample size. During the review process, we added number of exposure days as a covariate in the regression.

**Supplementary Tables**

**Supplementary Table 1: Characteristics of COVID-19 index cases by number of COVID-19 vaccine doses in California prisons**

|  | Index cases (N=1226) (N (%) or mean (SD)) | | | |
| --- | --- | --- | --- | --- |
|  | No COVID-19 vaccination (N=273) | 1 dose of COVID-19 vaccine (N=76) | 2 doses of COVID-19 vaccine (N=337) | >3 doses of COVID-19 vaccine  (N=540) |
| **Sex** |  |  |  |  |
| Female | 8 (3%) | 0 (0%) | 7 (4%) | 31 (3%) |
| Male | 265 (97.1) | 76 (100.0) | 327 (97.0) | 520 (96.3) |
| **Age (years)** | 36.3 (10) | 33.4 (7.8) | 37 (10.2) | 41.1 (10.8) |
| **Race** |  |  |  |  |
| American Indian/Alaskan Native | 0 (0%) | 1 (1%) | 2 (1%) | 8 (2%) |
| Asian or Pacific Islander | 4 (2%) | 1 (1%) | 5 (2%) | 6 (1%) |
| Black | 89 (33%) | 22 (29%) | 95 (28%) | 104 (19%) |
| Hispanic | 121 (44%) | 35 (46%) | 146 (43%) | 221 (41%) |
| Mexican | 24 (9%) | 6 (8%) | 48 (14%) | 92 (17%) |
| White | 28 (10%) | 10 (13%) | 32 (10%) | 94 (17%) |
| Other^*^ | 7 (3%) | 1 (1%) | 9 (3%) | 15 (3%) |
| **COVID-19 risk score (range: 0-12)^**^** | 0.7 (1.3) | 0.6 (0.8) | 0.8 (1%) | 1.2 (1.5) |
| **Number of days of exposure between index case and close contact** | 2.4 (1.2) | 2.43 (1) | 2.3 (1%) | 2.2 (1.1) |
| **Prior infection** | 84 (31%) | 33 (43%) | 123 (37%) | 200 (37%) |
| **Vaccination status** |  |  |  |  |
| *Unvaccinated* | 273 (100%) | - | - | - |
| *Ad26.COV2* | - | 58 (76%) | 55 (16%) | - |
| Completed only primary series | - | 58 (100%) | - | - |
| Received booster or additional doses | - | - | 55 (100%) | - |
| *BNT162b2* | - | 5 (6.6%) | 55 (16%) | 128 (24%) |
| Received 1 dose of primary series | - | 5 (100%) | - | - |
| Completed only primary series | - | - | 55 (100%) | - |
| Received booster or additional doses | - | - | - | 128 (100%) |
| *mRNA-1273* | - | 13 (17%) | 227 (67%) | 412 (76%) |
| Received 1 dose of primary series | - | 13 (100%) | - | - |
| Completed only primary series | - | - | 227 (100%) | - |
| Received booster or additional doses | - | - | - | 412 (100%) |

History of prior natural infection and vaccination status reflect the index case’s vaccination and natural infection status on the day of first positive test

*Other race/ethnicity based on self-reported data and those with mixed race/ethnicity

**COVID-19 risk score was estimated by California Correctional Health Care Services as weighted sum of different comorbidities most associated with severe COVID-19 complications

**Supplementary Table 2: Unadjusted estimates of the attack rate of SARS-CoV-2 infection among close contacts of index cases by COVID-19 vaccination and SARS-CoV-2 infection history of index cases**

| Attack rate (%) (95% CI) | No prior vaccination | Prior vaccination |
| --- | --- | --- |
| No prior infection | 39.2 (32.2, 46.5) | 31.5 (27.8, 35.4) |
| Prior infection | 29.8 (20.5, 40.9) | 21.3 (17.3, 26) |

**Supplementary Table 3: Primary analysis of the relationship of COVID-19 vaccination and prior SARS-CoV-2 infection on infectiousness of Omicron SARS-CoV-2 infections**

|  |  | Relative % change in attack rate of infection in close contact  (95% CI) |
| --- | --- | --- |
| Index case | Prior vaccination only | -22.4 (-36, -6) |
|  | Prior infection only | -22.6 (-38.5, -2.7) |
| Close contact | Duration of exposure (per day) | 6.9 (-2.3, 16.9) |
|  | Number of vaccine doses |  |
|  | 1 dose | 1.3 (-8.1, 11.8) |
|  | 2 doses | 2.7 (-15.5, 24.9) |
|  | ≥3 doses | 4.1 (-22.4, 39.6) |
|  | Prior infection only | -19.1 (-34.9, 0.6) |
| Institution | SARS-CoV-2 incidence in the 7 days preceding the positive test in the index case (per natural log increase in incidence) | 10.2 (-4.8, 27.6) |

The primary analysis estimated the relationship of the index cases’ vaccine status and prior natural infection history on attack risk of SARS-CoV-2 infection in the close contact. We adjusted for potential confounders, including the duration of exposure between index cases and close contacts, number of COVID-19 vaccine doses and prior natural infection history in close contacts as well as institution SARS-CoV-2 incidence.

**Supplementary Table 4: Testing alternative definitions of COVID-19 vaccination status in index cases and close contacts on the infectiousness of Omicron SARS-CoV-2 infections**

|  | COVID-19 vaccination status | Relative % change in attack rate of infection in close contact  (95% CI) |
| --- | --- | --- |
| Index case | Prior vaccination | -22.4 (-36, -6) |
|  | Number of vaccine doses |  |
|  | 1 dose | -11.4 (-17.4, -5.1) |
|  | 2 doses | -21.5 (-31.7, -9.9) |
|  | ≥3 doses | -30.5 (-43.5, -14.4) |
| Close contact | Prior vaccination | 2.3 (-21.8, 33.7) |
|  | Number of vaccine doses |  |
|  | 1 dose | 1.3 (-8.1, 11.8) |
|  | 2 doses | 2.7 (-15.5, 24.9) |
|  | ≥3 doses | 4.1 (-22.4, 39.6) |
|  | Prior vaccination or infection | -10.5 (-33.4, 20.2) |

This sensitivity analysis represents distinct regression analyses measuring the infectiousness of SARS-CoV-2 infections in index cases among close contacts under different definitions of COVID-19 vaccine status in the index case and close contact. The sensitivity analysis was conducted by starting with the primary model and making a single change to either the index case or close contact’s vaccine status under three definitions: (1) any vaccination as a binary variable; (2) vaccination as number of doses; (3) combined any vaccination and/or prior infection as binary definition. Weights for vaccinated index cases were rebalanced for the analysis defining vaccine status in index cases as number of doses.

**Supplementary Table 5: Sensitivity analysis matching vaccinated index cases by number of doses to unvaccinated index cases to examine relationship between number of COVID-19 vaccine doses and infectiousness of Omicron SARS-CoV-2 infections**

| Index case number of vaccine doses | Relative % change in attack rate of infection in close contact (95% CI) |
| --- | --- |
| 1 dose | -12.4 (-18.2, -6.2) |
| 2 doses | -23.2 (-33.1, -12) |
| ≥3 doses | -32.8 (-45.2, -17.5) |

To understand the relationship between the number of vaccine doses in an index case and their infectiousness, we reweighted matches from the primary analysis to reflect three separate vaccine groups (1 dose, 2 dose, 3 doses). In this sensitivity analysis, we matched unvaccinated cases with the three vaccinated groups with 1:10 matching. All other aspects of the analysis remained the same.

**Supplementary Table 6: Assessment of statistical interaction between COVID-19 vaccination and prior SARS-CoV-2 infection in the index case on the infectiousness of Omicron SARS-CoV-2 infections**

| Index case | Relative % change in attack rate of infection in close contact (95% CI) | |
| --- | --- | --- |
|  | Without interaction term | With interaction term |
| Prior vaccination | -22.4 (-36, -6) | -22.6 (-37.7, -3.9) |
| Prior infection | -22.6 (-38.5, -2.7) | -23.2 (-47.9, 13.4) |
| Interaction between vaccination and prior infection | -- | 0.9 (-36.2, 59.7) |

**Supplementary Table 7: Estimating the relationship between time since most recent COVID-19 vaccine dose or natural infection on infectiousness of Omicron SARS-CoV-2 infections**

|  | Relative % change in attack rate of infection in close contact (95% CI) |
| --- | --- |
| Time since last COVID-19 vaccine dose (per 5 weeks) | 6.4 (2.3, 10.6) |
| Time since most recent SARS-CoV-2 infection (per 5 weeks) | 4.7 (-3.6, 13.7) |
| Time since most recent vaccine or infection (per 5 weeks) | 4.7 (1.9, 7.6) |

We conducted three regression analyses estimating the relationship between: 1) time since most recent vaccination and risk of transmission of infection; 2) time since most recent infection and risk of transmission of infection; 3) time since most recent vaccination or infection and risk of transmission of infection. All analyses were adjusted for prior vaccination and/or infection in the index case as well as close contact and institution-specific characteristics. Of note, these analyses assume a SARS-CoV-2 infection (whereas immunity from vaccine and/or natural infection may prevent infection soon after vaccine and/or natural infection) thus limiting the ability to detect a relationship.

**Supplementary Table 8: Sensitivity analysis on inclusion and exclusion criteria of the primary study design**

|  | Index case | Relative % change in attack rate of infection in close contact  (95% CI) |
| --- | --- | --- |
| Removing requirement for negative test in close contact within 2 days of first exposure | Prior vaccination | -19 (-32.7, -2.6) |
|  | Prior infection | -20.7 (-36.7, -0.6) |
| Including close contacts that test positive within 2 days after first exposure | Prior vaccination | -23 (-35.4, -8.3) |
|  | Prior infection | -17.4 (-33.7, 2.9) |

**Supplementary Table 9: Sensitivity analysis testing alternative matching specifications for primary analysis on the infectiousness of Omicron SARS-CoV-2 infections**

| Matching specification | | Index case | Relative % change in attack rate of infection in close contact (95% CI) |
| --- | --- | --- | --- |
| Varying choice of caliper | Caliper of propensity score = 0.2 | Prior vaccination | -21 (-35.2, -3.8) |
|  |  | Prior infection | -25.3 (-41.3, -5.1) |
|  | Caliper of days between index cases = 15 | Prior vaccination | -23.4 (-36.7, -7.4) |
|  |  | Prior infection | -21.9 (-38, -1.6) |
| Varying k in 1:k matching | 1:4 matching | Prior vaccination | -21.2 (-35.2, -4.2) |
|  |  | Prior infection | -21.8 (-38.6, -0.5) |
| Varying weights between propensity score and time | 1:3 ratio | Prior vaccination | -21 (-34.9, -4) |
|  |  | Prior infection | -22.1 (-38, -2.2) |
|  | 3:1 ratio | Prior vaccination | -23.5 (-36.8, -7.3) |
|  |  | Prior infection | -24.5 (-40.7, -3.7) |
| Overall changes to propensity score | Matching without propensity score | Prior vaccination | -22.1 (-35.5, -5.9) |
|  |  | Prior infection | -23.6 (-39.7, -3.1) |
|  | Propensity score with sex and race | Prior vaccination | -24.2 (-36.9, -8.9) |
|  |  | Prior infection | -22.7 (-37.6, -4.1) |
|  | No matching | Prior vaccination | -23 (-36.7, -6.4) |
|  |  | Prior infection | -23 (-39.8, -1.5) |

**Supplementary Table 10: Sensitivity analysis testing alternative definitions of infectious period in index case for primary analysis on the infectiousness of Omicron SARS-CoV-2 infections**

| Start of infectious period | Duration of infectious period | Index case | Relative % change in attack rate of infection in close contact (95% CI) |
| --- | --- | --- | --- |
| Begins day of first positive test | 7 days | Prior vaccination | -20.3 (-34.7, -2.9) |
|  |  | Prior infection | -20.7 (-37.5, 0.6) |
| Begins 2 days prior to first positive test | 5 days | Prior vaccination | -13.9 (-28.4, 3.7) |
|  |  | Prior infection | -22.7 (-38.9, -2.2) |
|  | 7 days | Prior vaccination | -12.5 (-26.9, 4.7) |
|  |  | Prior infection | -21 (-37.7, 0.2) |

**Supplementary Table 11: Sensitivity analysis assessing impact of excluding index cases that received the *Ad26.COV2* vaccine**

| Index case | Relative % change in attack rate of infection in close contact (95% CI) |
| --- | --- |
| Prior vaccination | -21.6 (-35, -5.4) |
| Prior infection | -19.1 (-36.9, 3.8) |

**Supplementary Table 12: Sensitivity analysis using logistic regression model to evaluate the infectiousness of Omicron SARS-CoV-2 infections**

| Index case | Odds ratio of infection in close contact (95% CI) |
| --- | --- |
| Prior vaccination | 0.66 (0.48, 0.91) |
| Prior infection | 0.68 (0.49, 0.95) |

Note: The primary analysis used a robust Poisson regression model, while this sensitivity

analysis used a logistic regression model.

**Supplementary Table 13: Estimation of the fraction of secondary transmission attributable to Omicron SARS-CoV-2 infections stratified by prior COVID-19 vaccination and natural infection history**

|  | Fraction of transmission from SARS-CoV-2 infections (%) (95% CI) | | |
| --- | --- | --- | --- |
|  | Within secondary cases in study population | Among all Omicron SARS-CoV-2 infections | Among Omicron SARS-CoV-2 infections with complete prior infection history |
| No prior vaccination or infection | 20.4 (16.4, 25) | 23.5 (27.7, 19.8) | 14.9 (18, 12.2) |
| Prior vaccination only | 51.8 (46.5, 57) | 52.8 (51.2, 53.8) | 48.2 (48.1, 47.8) |
| Prior infection only | 6.9 (4.6, 10.1) | 4.4 (4.1, 4.7) | 6.6 (6.3, 6.8) |
| Both prior vaccination and infection | 20.9 (16.9, 25.6) | 19.3 (17, 21.6) | 30.4 (27.6, 33.2) |

We estimated the attributable fraction of transmission from SARS-CoV-2 infection among secondary cases, stratified by different vaccine and immune statuses of the index case. We performed this analysis under three circumstances: (1) study population; (2) among all confirmed SARS-CoV-2 infections in the study period; and (3) among confirmed SARS-CoV-2 infections in residents that were incarcerated before April 2020 due to underreporting of prior infection history. Further description is available in the main text.

**Supplementary Table 14. Characteristics of included and excluded close contacts.**

|  | Included close contacts (N=1388) (N (%) or mean (SD)) | Excluded close contacts (N=1171) (N (%) or mean (SD)) |
| --- | --- | --- |
| **Sex** |  |  |
| Female | 41 (3%) | 1 (0%) |
| Male | 1347 (97%) | 1170 (100%) |
| **Age (years)** | 40.2 (11.8) | 39.4 (11) |
| **Race/ethnicity** |  |  |
| American Indian/Alaskan Native | 14 (1%) | 11 (1%) |
| Asian or Pacific Islander | 10 (1%) | 16 (1%) |
| Black | 351 (25%) | 362 (31%) |
| Hispanic | 750 (54%) | 573 (49%) |
| White | 215 (16%) | 172 (15%) |
| Other^*^ | 48 (4%) | 37 (3%) |
| **COVID-19 risk score (range 0-12)^**^** | 1.2 (1.6) | 1 (1.5) |
| **Prior infection** | 565 (41%) | 419 (36%) |
| **Vaccination status** |  |  |
| *Unvaccinated* | 197 (14%) | 220 (19%) |
| *Ad26.COV2* | 159 (11%) | 112 (9.6%) |
| Completed only primary series | 73 (46%) | 46 (41%) |
| Received booster or additional doses | 86 (54%) | 66 (59%) |
| *BNT162b2* | 246 (18%) | 176 (15%) |
| Received 1 dose of primary series | 3 (1.2%) | 2 (1.1%) |
| Completed only primary series | 49 (20%) | 40 (23%) |
| Received booster or additional doses | 194 (79%) | 134 (76%) |
| *mRNA-1273* | 786 (57%) | 663 (57%) |
| Received 1 dose of primary series | 28 (3.6%) | 20 (3%) |
| Completed only primary series | 239 (30%) | 220 (33%) |
| Received booster or additional doses | 519 (66%) | 423 (64%) |

Contacts were included if they met all study criteria. Excluded close contacts include close contacts that met study criteria but are missing either a negative test within 2 days of exposure to an index case and/or follow up testing data. If residents were considered close contacts for multiple index cases, we randomly assigned an index case for these purposes. We excluded any residents that were valid close contacts for an index case(s) and excluded for another index case(s). History of prior natural infection and vaccination status reflect the close contact’s vaccination and natural infection status on the day of first exposure to an index case.

*Other race/ethnicity based on self-reported data and those with mixed race/ethnicity

**COVID-19 risk score was estimated by California Correctional Health Care Services as weighted sum of different comorbidities most associated with severe COVID-19 complications

**Supplementary Figures**

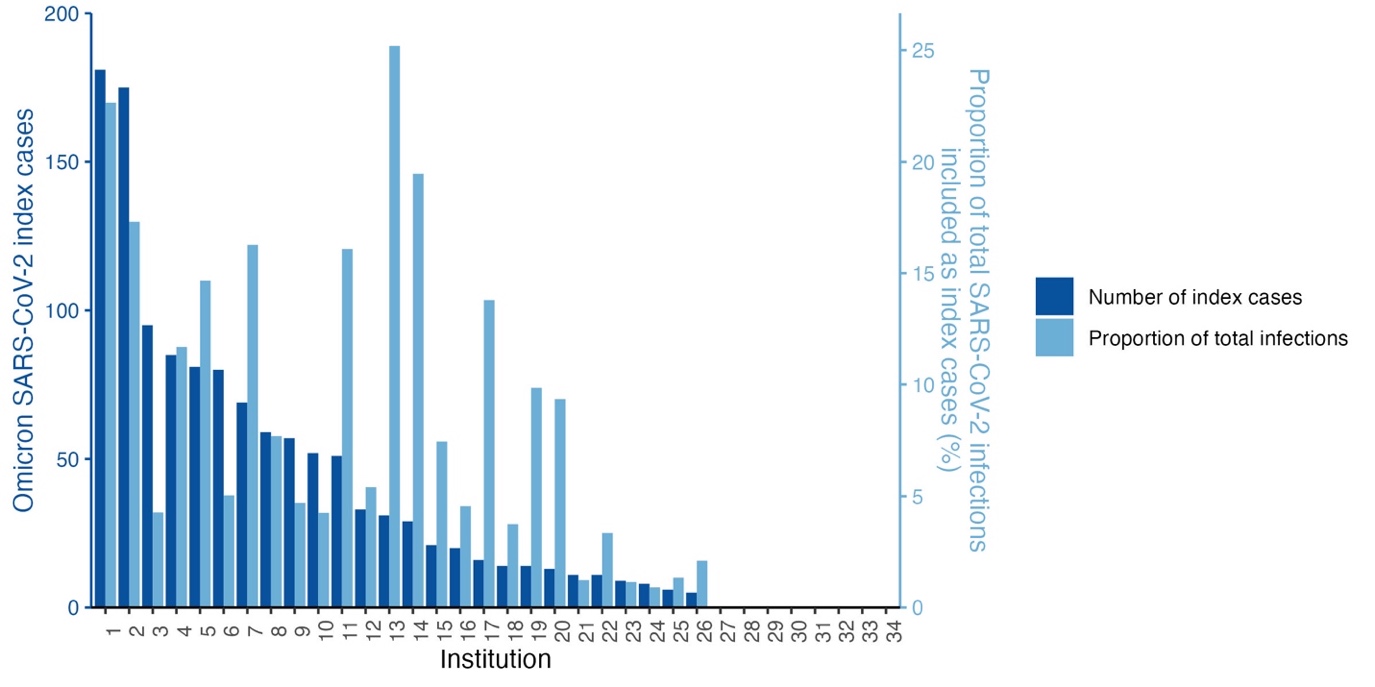

**Supplementary Figure 1: Omicron SARS-CoV-2 infections included as index cases by institution**. We plotted the absolute number of index cases included in the analysis by institution (dark blue) with y-axis on left. We plotted the proportion of total Omicron SARS-CoV-2 infections included as index cases by institution (light blue) with y-axis on right. Infections from some institutions are overrepresented in our final sample due to our strict inclusion and exclusion criteria established to address confounding and misattribution concerns.

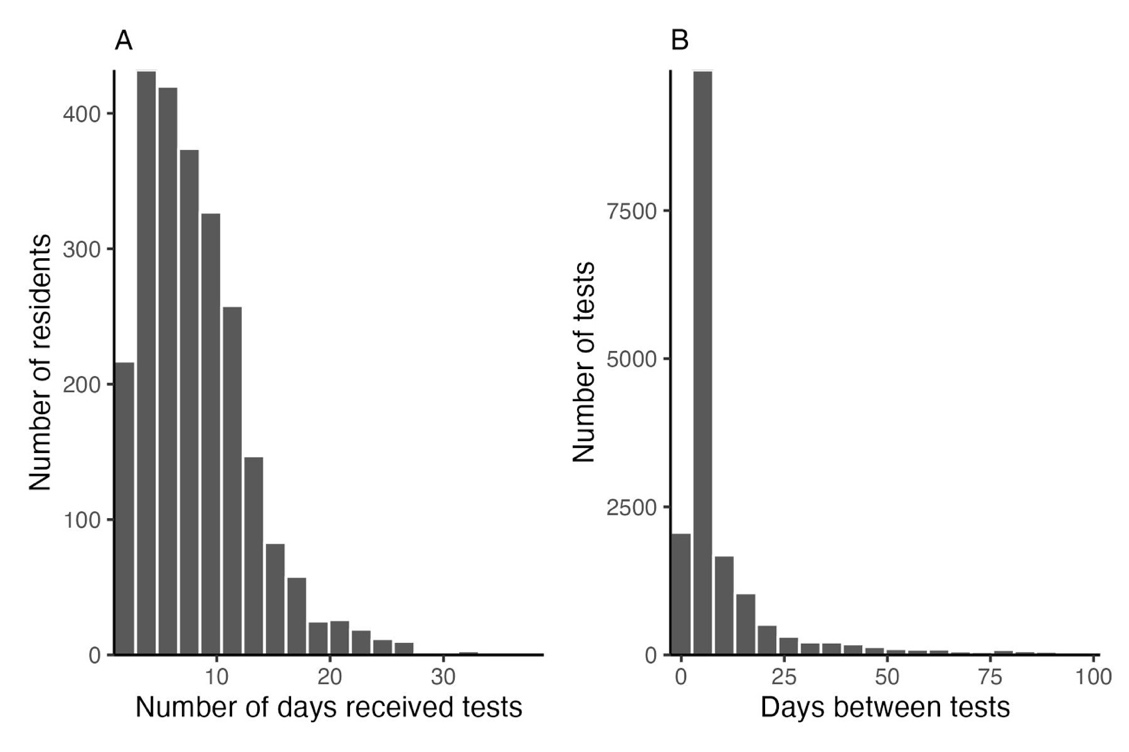

**Supplementary Figure 2: Number of and time between SARS-CoV-2 tests.** Here we plot the distributions of measures of testing frequency in the study population over the study period (December 15, 2021 – May 20, 2022).

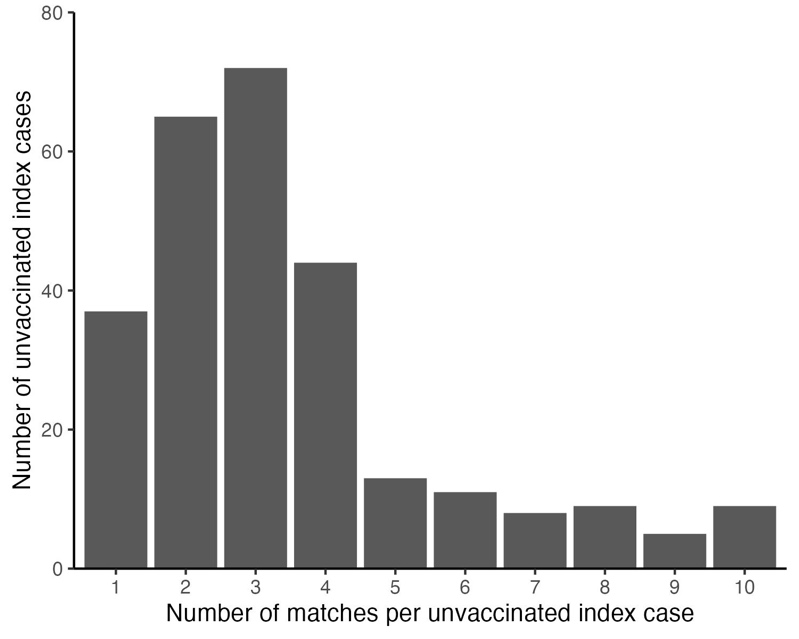

**Supplementary Figure 3:** **Number of vaccinated index case matches for unvaccinated index cases.** We performed 1:10 matching of unvaccinated (N=276) and vaccinated (N=953) COVID-19 index cases and plot the distribution of the number of matches for unvaccinated COVID-19 cases.

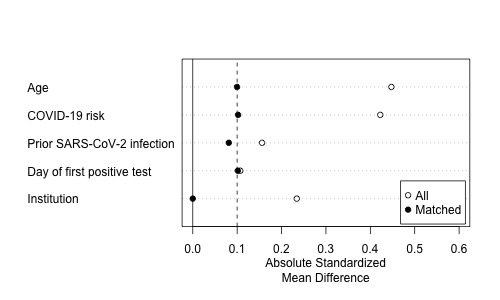

**Supplementary Figure 4: Balance across matching variables before and after matching.** We performed 1:10 matching of unvaccinated (N=273) and vaccinated (N=953) COVID-19 index cases by institution, day, and propensity for vaccination (based on age, COVID-19 risk, and prior SARS-CoV-2 infection). We plotted the standardized mean differences between index cases before and after matching. The dashed line at 0.1 represents a recommended threshold for differences between exposure groups when matching.

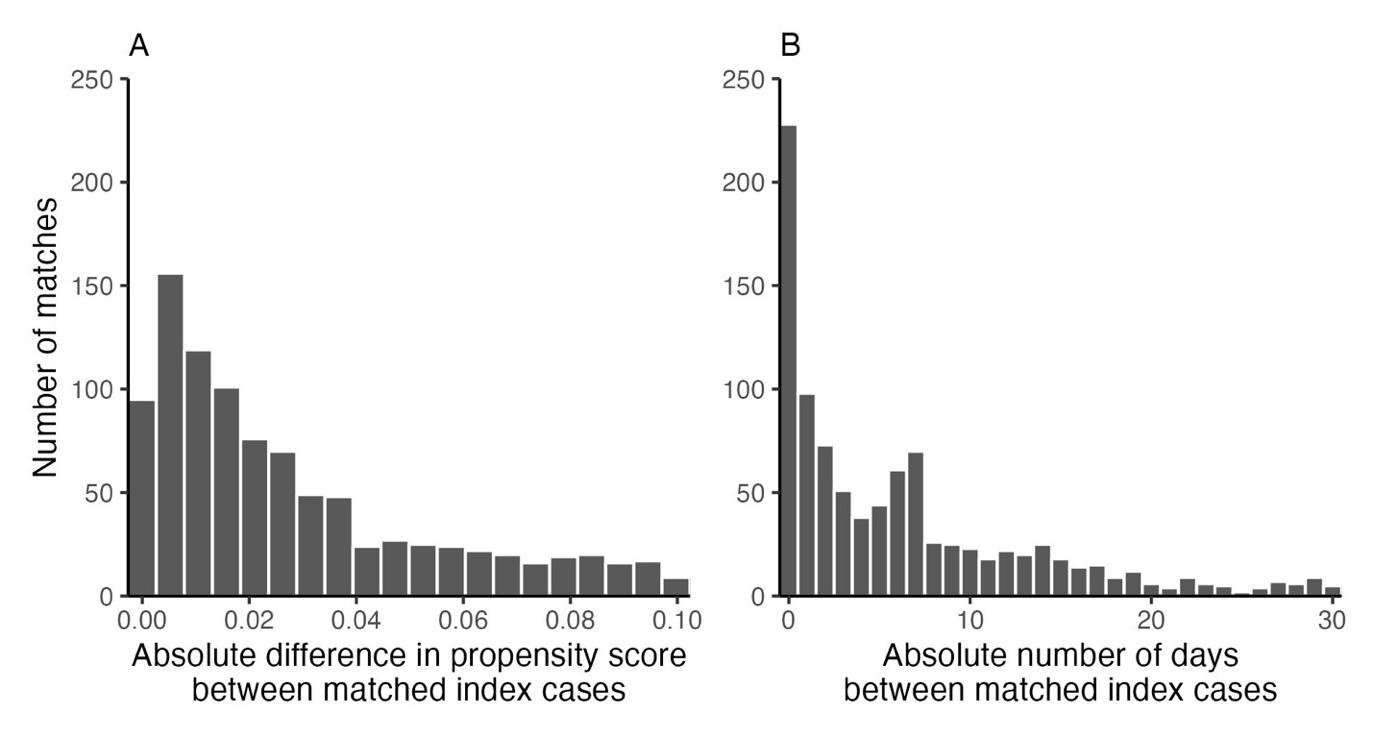

**Supplementary Figure 5: Distance between matched unvaccinated and vaccinated index cases.** We performed 1:10 matching of unvaccinated (N=273) and vaccinated (N=953) COVID-19 index cases by institution, day (caliper = 30 days), and propensity for vaccination (based on age, COVID-19 risk, and prior SARS-CoV-2 infection, caliper = 0.1). We plot the absolute differences in propensity score (A) and days (B) between matches.

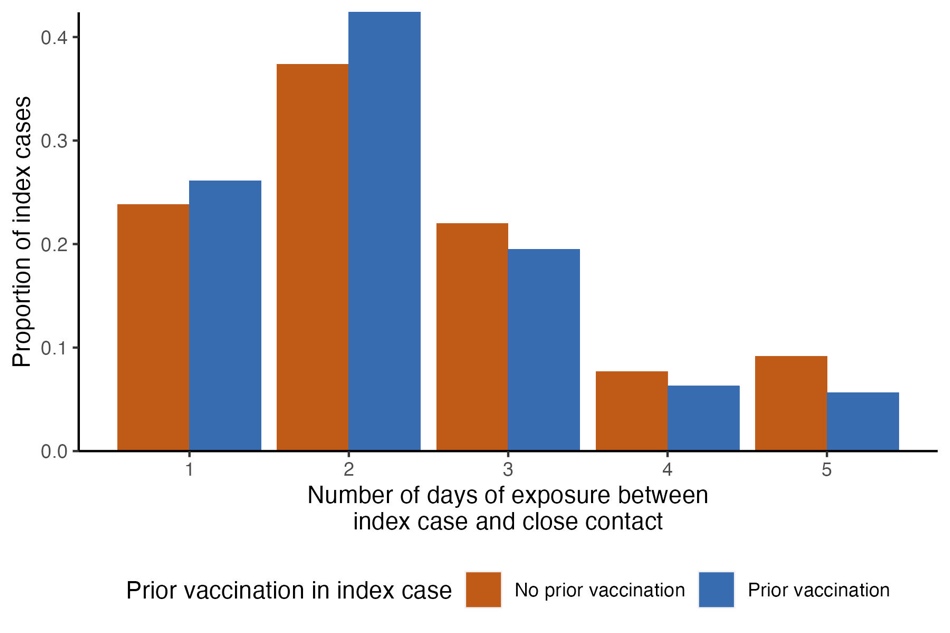

**Supplementary Figure 6: Exposure between the index case and close contact during the infectious period.** Given residents were frequently moved, including for isolation, we plotted the number of days of close contact with the index case during their infectious period for the study period, stratified by the index case’s vaccination status.

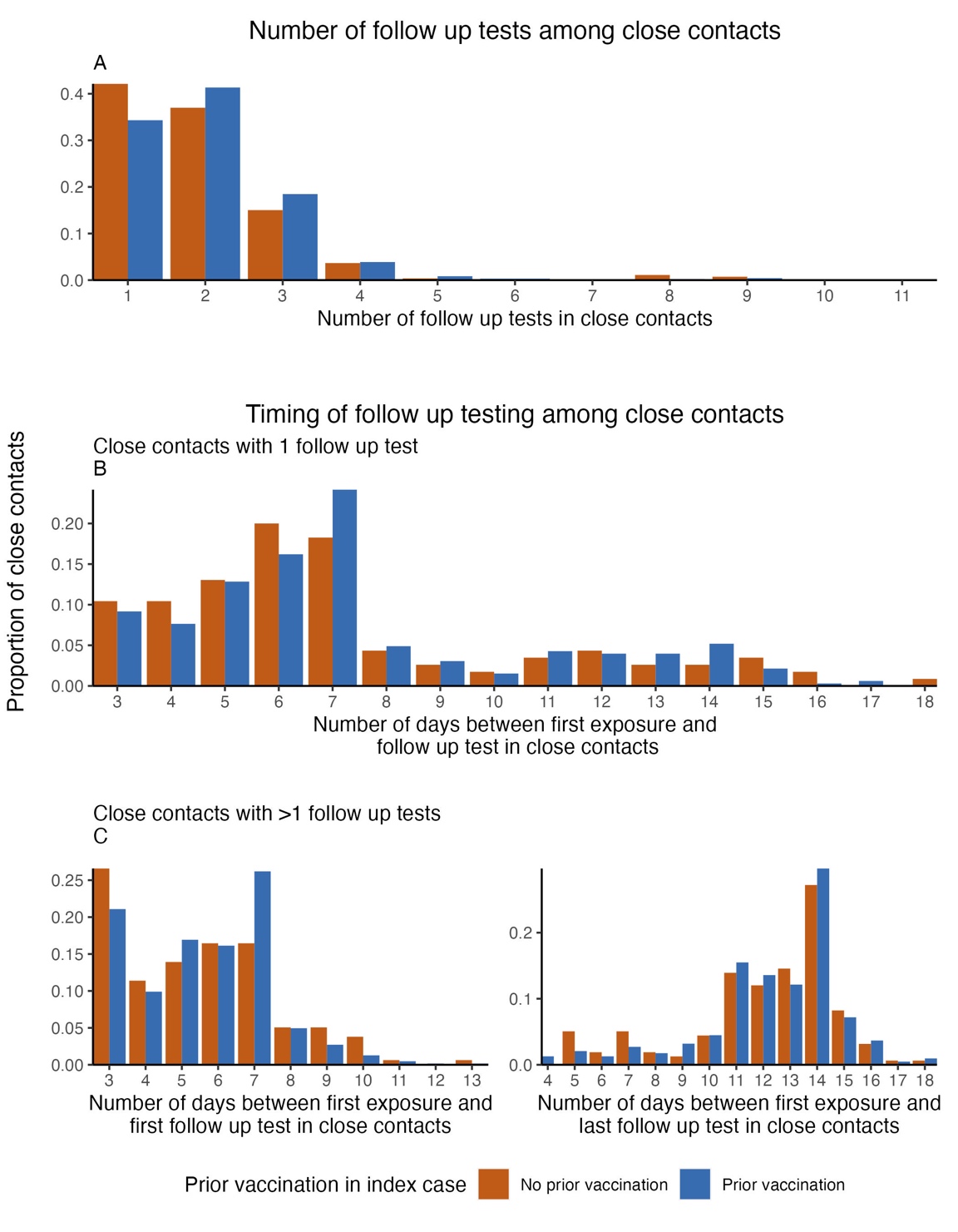

**Supplementary Figure 7: Follow up testing in close contacts after exposure to index case.** Distribution of the number of follow up tests and the timing of testing among close contacts in the study population

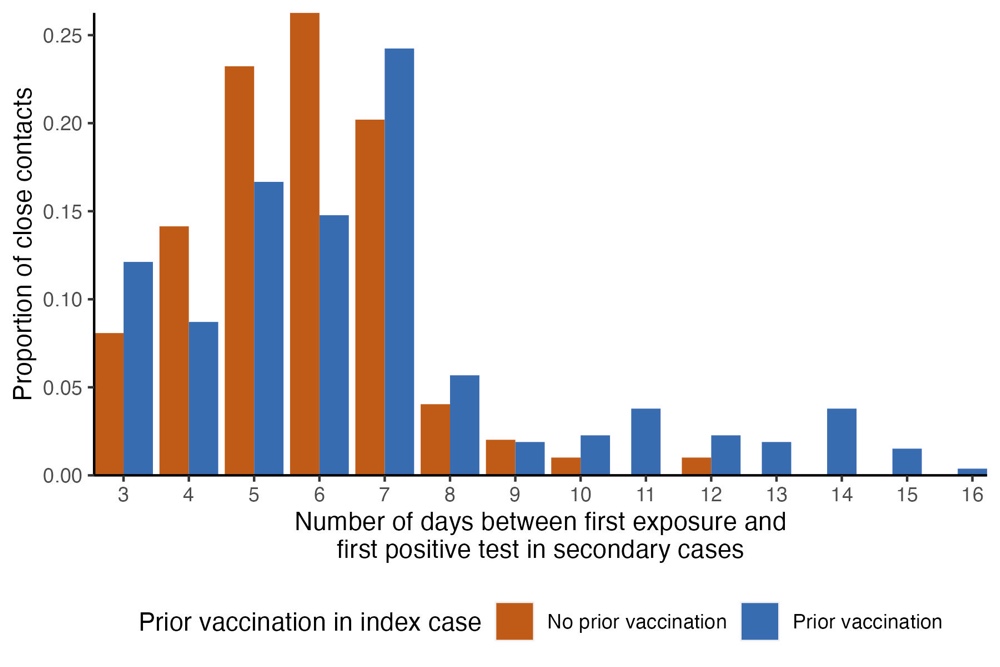

**Supplementary Figure 8: Timing of secondary SARS-CoV-2 infections.** We defined secondary SARS-CoV-2 infections as close contacts who tested positive for SARS-CoV-2 during the follow up period. We plot the distribution of the number of days between a secondary case’s first exposure to an index case and their first positive test.

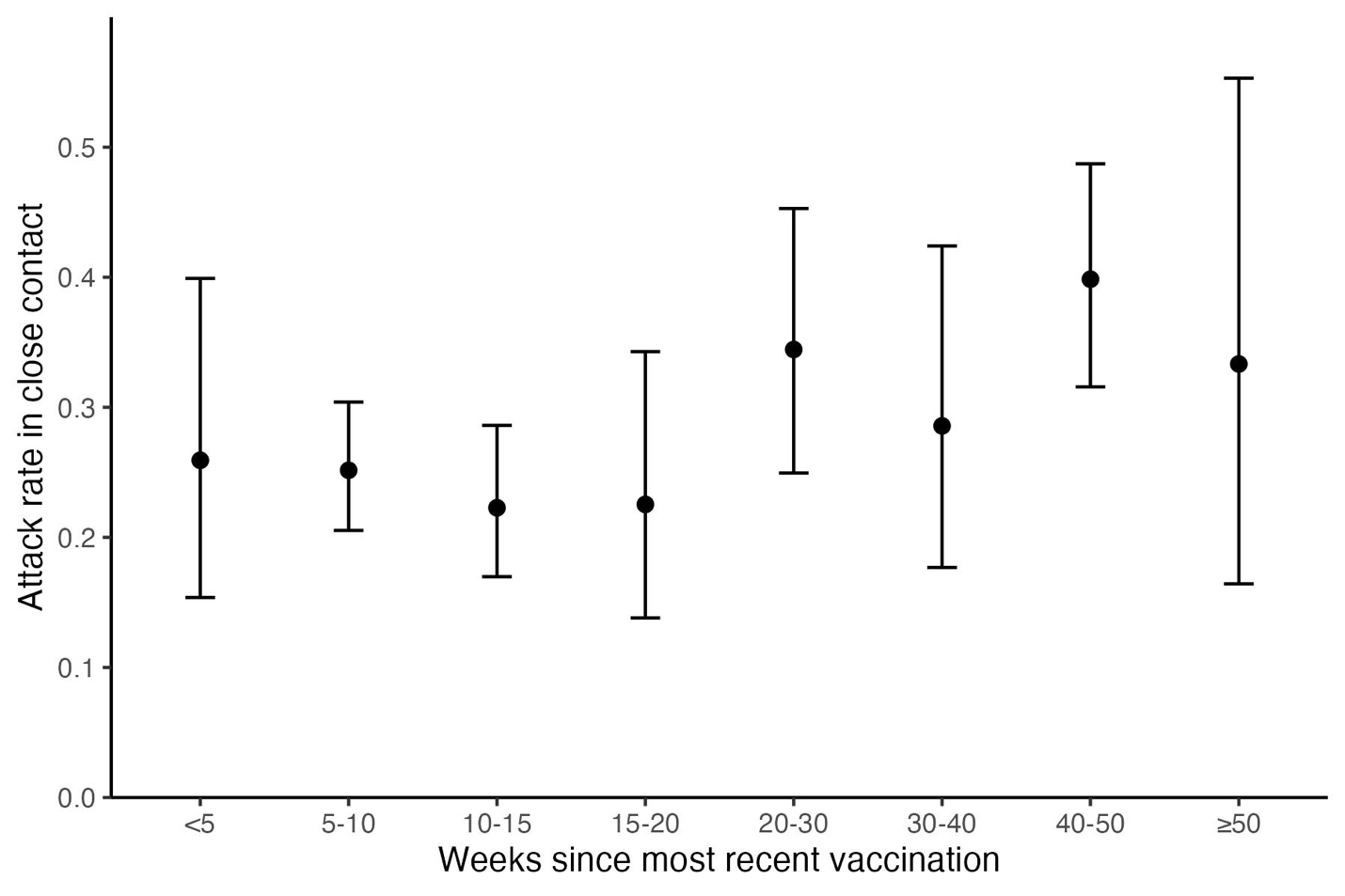

**Supplementary Figure 9: Unadjusted estimates of the attack rate of Omicron SARS-CoV-2 infection of vaccinated index cases by time since the index cases’ most recent vaccine dose.** We plotted the unadjusted attack rate (represented by points) and 95% binomial confidence intervals (represented by error bars) for vaccinated index cases, stratified by time (in weeks) since the index cases’ most recent vaccine dose prior to first positive SARS-CoV-2 test. The adjusted estimates from the regression model are available in Supplementary Table 5.
